# Sensorimotor cortex forms prism adaptation memories in older adults and stroke patients

**DOI:** 10.1101/2020.12.02.20242297

**Authors:** Gershon Spitz, Pierre Petitet, Janet Bultitude, Alessandro Farnè, Jacques Luaute, Jacinta O’Shea

**Affiliations:** Wellcome Centre for Integrative Neuroimaging, Nuffield Department of Clinical Neurosciences, Oxford, United Kingdom; Turner Institute for Brain and Mental Health, Monash University, Melbourne, Australia; Centre for Pain Research, University of Bath, Bath, United Kingdom; Department of Psychology, University of Bath, Bath, United Kingdom; Integrative Multisensory Perception Action Cognition Team - ImpAct, Lyon Neuroscience Research Center, INSERM U1028, CNRS U5292, Lyon, France; Rehabilitation department, Hospices Civils de Lyon, Henry Gabrielle, Hospital, Saint Genis Laval, France; University of Lyon 1, Lyon, France; Hospices Civils de Lyon, Mouvement Handicap, Neuro-immersion, Lyon, France; Center for Mind/Brain Sciences, University of Trento, Rovereto, Italy; Wellcome Centre for Integrative Neuroimaging, Oxford Centre for Human Brain Activity (OHBA),University of Oxford Department of Psychiatry, Warneford Hospital, Warneford Lane, Oxford, United Kingdom

**Keywords:** prism adaptation, stroke, neglect, spatial attention, computational neuroimaging

## Abstract

Stroke is the largest cause of complex disability in adults. Approximately half of right-hemisphere stroke survivors suffer spatial neglect–an inability to voluntarily orient to people or objects in contralesional space. Neglect is a significant impediment to successful community reintegration. Prism adaptation (PA) is a promising behavioural intervention that can alleviate symptoms of spatial neglect. PA induces a leftward pointing bias–the prism after-effect (AE). In neglect, the prism AE generalises to improve other sensory, motor, and cognitive domains. Although the formation of an AE is a key index in neglect, we do not yet know where it is formed in the brain. Here, we used a novel computational fMRI-based approach to study, for the first time, the brain circuits that mediate the formation of PA in stroke survivors and age matched controls. Healthy individuals (*n* = 17) and stroke patients (*n* = 11) performed prism adaptation during fMRI. Temporal signatures of memory formation were extracted from the behavioural data using a state-space model and regressed against the fMRI data. This revealed that, in both groups, fMRI signal in left sensorimotor cortex correlated with the gradual formation of the prism after-effect during adaptation. This indicates that the sensorimotor cortex may be a useful target for neuromodulation that aims to improve the persistence of therapeutic prism after effects.

## 1. Introduction

Stroke is the largest cause of complex disability in adults (Adamson et al., 2004). Half of right-hemisphere stroke survivors experience spatial neglect– characterised by a failure to orient voluntarily to people or objects in contralesional space (Stone et al., 1993, Ringman et al., 2004). Spatial neglect is a significant source of long-term disability, resulting in poorer engagement with occupational, social, and leisure activities (Di Monaco et al., 2011, Katz et al., 1999). Despite spatial neglect being a profound source of disability, there is limited evidence that current rehabilitation approaches lead to consistent and sustained improvements in function (Bowen et al., 2013, Azouvi et al., 2017).

Prism adaptation (PA) is a promising behavioural paradigm that can reduce the symptoms of spatial neglect under controlled experimental conditions (Rossetti et al., 1998, O’Shea et al., 2017). Glasses fitted with prismatic lenses are used to induce a rightward shift in the visual field causing individuals to make rightward errors during pointing movements to visual targets. Individuals learn to correct these errors through feedback from successive pointing movements. Subsequent removal of the prism glasses results in aleftward pointing bias – the prism after-effect. In neglect, this leftward bias generalises to improve other sensory, motor, and cognitive impairments in neglect (Jacquin-Courtois et al., 2013). This generalisation makes prism adaptation one of the more promising treatments for spatial neglect.

However, these prism after effects are short-lasting. While a single session of prism adaptation can induce improvements that last for up to a few days (Rossetti et al., 1998, Farnè et al., 2002, Pisella et al., 2002, Jacquin-Courtois et al., 2008), repeated sessions are required to prolong benefit (Serino et al., 2006), while others have not found any long-term beneficial effects (Rousseaux et al., 2006, Rode et al., 2015). We, and others, have argued that the efficacy of prism therapy for neglect could be improved through better characterisation, and subsequent targeted neuromodulation, of the key brain circuitry that drives prism adaptation behaviour, specifically the formation and persistence of the prism after effect (Barrett et al., 2012, Petitet et al., 2017, O’Shea et al., 2017).

A recent review of the literature on the neural bases of prism adaptation (Panico et al., 2019). concluded that: 1. cerebellar-parietal networks mediate the sensorimotor aspects of prism adaptation, 2. temporal and prefrontal regions mediate generalization to other cognitive domains, and 3. that formation and retention of the prism after-effect depends on the motor cortex. The latter arose from our previous work in which we showed that stimulating left sensorimotor cortex during adaptation caused longer lasting sensorimotor and cognitive prism after-effects in healthy volunteers and neglect patients (O’Shea et al., 2017).

Previous neuroimaging studies of adaptation have focused on localizing processes of error correction. Here, instead, we aimed to identify the neural circuits that mediate the formation of the prism after-effect. To identify brain signals associated with the gradual development of the prism effect during adaptation, we fitted a state-space model to the behavioural data. Adaptation is well-described by state space models that account for behaviour as the output of two or more parallel learning systems operating on different timescales, fast and slow (Smith et al., 2006, Kording et al., 2007, Joiner and Smith, 2008, Kim et al., 2015). We developed a state space model variant that captured the gradually changing magnitude and stability of the prism after effect during adaptation (Petitet et al., 2018). We used this to identify functional brain signals whose timecourse correlated with the fast and slow learning dynamics that underpin the formation of the behavioural after effect.

Healthy individuals and stroke patients performed prism adaptation during fMRI (Bultitude et al., 2016). Using a similar approach to Kim et al. (2015), a computational model was fitted to prism adaptation behaviour to extract time-varying regressors of brain activity that follow a fast or slow time-scale (Petitet et al., 2018). In both healthy individuals and stroke patients, functional signal in left sensorimotor cortex correlated with the slow timescale dynamics, indicating that this brain region mediates the formation of the prism after effect.

## 2. Materials and Methods

Description of the study apparatus construction and evaluation, participant samples, MRI acquisition, and initial behavioural results are published else-where (Bultitude et al., 2016).

### 2.1. Participants

The study was approved according to **\*omitted text for preprint**. All Healthy subjects and patients provided written informed consent. Eighteen healthy controls and 12 stroke patients volunteered for the current study. All participants were right-hand dominant. Stroke patients suffered a first-time right-hemisphere stroke and displayed symptoms of spatial neglect at some point following their injury (see Appendix .1 for lesion overly figure). At the time of testing only two stroke patients presented with symptoms of neglect. This was defined as at least two target omissions on the left of the page compared to the right on the Bell’s cancellation test (Azouvi et al., 2002). Stroke patients and healthy controls had no other history of neurological injury or illness. One stroke patient was excluded due to a piece of occluding material becoming dislodged from the head of the MRI coil, preventing the participant from seeing all task targets. One healthy control participant was excluded due to discomfort during the session, making them unable to reach the leftmost task target. The final study sample comprised 17 healthy controls (mean age = 61, SD = 13.9) and 11 stroke patients (mean age = 52, SD = 10.4, Table 1).

**Table 1:**
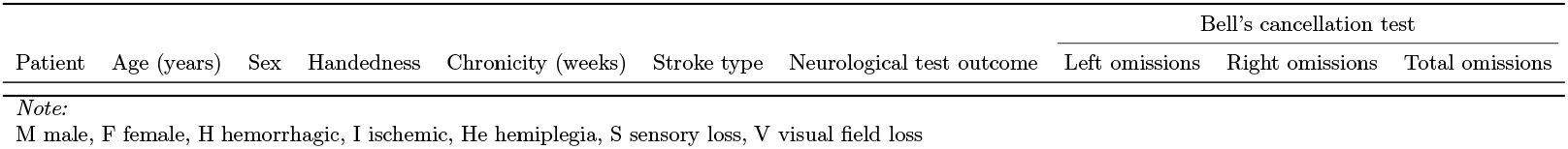
Patient demographics.

*Detailed participant characteristics in Table 1 have been omitted in preprint to reduce identifying information.

### 2.2. Behavioural protocol

Participants completed a prism adaptation behavioural paradigm while in the MRI scanner. The protocol comprised two consecutive fMRI runs: a) a sham run, in which participants made pointing movements towards an LED target on a screen, seen through a neutral, clear, Perspex sheet, and b) a real prism run, in which participants made pointing movements towards a target viewed through a Fresnel sheet that applied a 7.7^*°*^ prismatic shift to the visual field.

Each run comprised 20 behavioural blocks, each consisting of six pointing trials. Each sham and prism run comprised a total of 120 pointing trials. The inter-trial interval was 6.5 ±1sec and inter-block interval was 7.5 ± 1sec. Pointing blocks alternated between blocks of prism exposure, in which participants learned from feedback to correct their errors, and blocks of after-effect measurement, in which no visual feedback was provided. During prism exposure blocks, participants made rapid reaching movements towards an LED target located at either left or right positions on a screen located in front of them. Visual feedback was limited to the second portion of movement to minimise the effect of ‘online’ corrections on pointing trajectory. During after-effect trials, participants reached and pointed at an LED target at the center of the screen, viewed through the neutral Perspex sheet. Visual feedback was occluded via a panel that was lowered at the initiation of each pointing movement.

### 2.3. MRI acquisition protocol

Neuroimaging data were acquired at **\*omitted text for preprint**, France, on a Siemens Magnetom Sonata 1.5T scanner with a circularly polarized head coil with two integrated preamplifiers. Anatomical images were acquired with a MPRT1 sequence (176 slices, FOV = 256 mm, TR/TE/flip angle = 1.97/3.93/15^*°*^, 1.0 × 1.0 × 1.0 mm voxels) taking 8.26 minutes. Functional MRI of the whole brain was acquired in the axial plane (26 slices, FOV = 220 × 220mm^2^, *TR/TE/flipangle* = 2, 500*ms/*60*ms/*90, 3.4 × 3.4 × 5.0 mm voxels). Each of the two functional runs lasted 12.35 min.

### 2.4. Data analysis

#### 2.4.1. Modelling pointing accuracy

Reach endpoint errors were measured as the lateral deviation relative to the LED screen targets (measured in pixels, Px), which formed the behavioural measure of pointing accuracy. During the *real prism* run, rightward pointing errors were expected at prism onset, which would gradually be corrected across trials until baseline accuracy was restored. Interleaved between blocks of prism exposure the after effect was measured as a leftward pointing error that would gradually stabilize over time. A linear mixed-effect model was used to test for this expected behavioural profile. The analysis compared behaviour in the real versus sham prism runs and had the following factors: trial number, session (sham vs prism), group (control vs stroke), and a session × group interaction term. Participant (random intercept) and trial number (Random slope) were entered as random factors. Statistical analyses of behaviour were performed using R (R Core Team, 2017) in conjunction with the the lme4 package (Bates et al., 2015).

#### 2.4.2 Weighted two-state model of prism adaptation

A linear time-invariant weighted two-state model was used to estimate the contribution of fast and slow learning dynamics to the pointing behaviour during adaptation (see Petitet et al. (2018) for full characterisation and validation of this model). The model describes adaptation behaviour as the summed output of two parallel interacting systems that each learn and forget at different rates - one *fast* one *slow* system. The rapid error correction that occurs early during prism exposure is dominated by the fast system, whereas the stable pointing behaviour when adaptation has reached asymptote is primarily accounted for by the slow system. On error correction trials during prism exposure, the model is defined as follows:

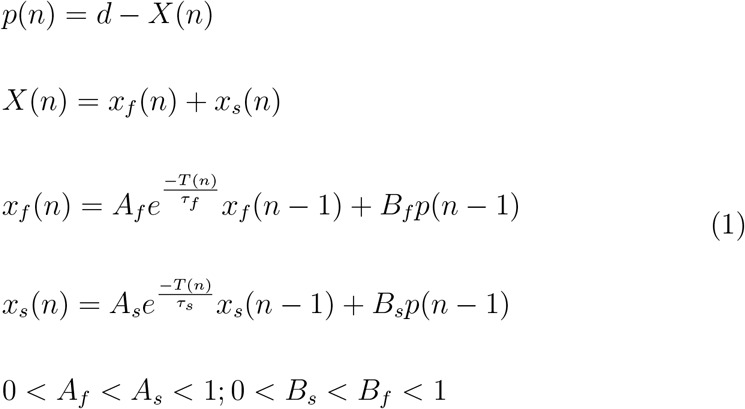

where the endpoint error on the n-th trial *p*(*n*) is modelled as the direct effect of the prism *d* minus the sum of the state of the two systems *X*(*n*). The direct effect *d* is a fixed parameter and is the group-averaged error magnitude on the first trial of prism exposure. It therefore represents the maximum rightward pointing error that the systems learn to gradually correct over time. Owing to online error corrections during the reach trajectory, *d* is typically 60-80% of the 7.7^*°*^ optical displacement. On any given trial the total amount of adaptation *X*(*n*) corresponds to the sum of the states of the fast system *x*_*f*_ (*n*) and the slow system *x*_*s*_(*n*). These systems produce a trial-by-trial estimate of *d* based on pointing error feedback. The learning rate *B* dictates the proportion of error added to the state of the systems from one trial to the next. Larger *B* means faster learning, i.e. a larger proportion of error on the (*n* − 1) − *th* trial is corrected for on trial n. Time decay (i.e. proportion of state that is remembered from one time point to the next) is modelled as an exponential decay function of time where A is an intercept (starting point), *τ* is a time constant and *T* (*n*) is the inter-trial interval between trial *n-1* and *n*.

On prism after-effect trials, *d, B*_*f*_ and *B*_*s*_ are set to zero to reflect the removal of the prismatic shift and the absence of visual feedback of the endpoint error.

In addition, weighting factors are introduced for each system as follows:

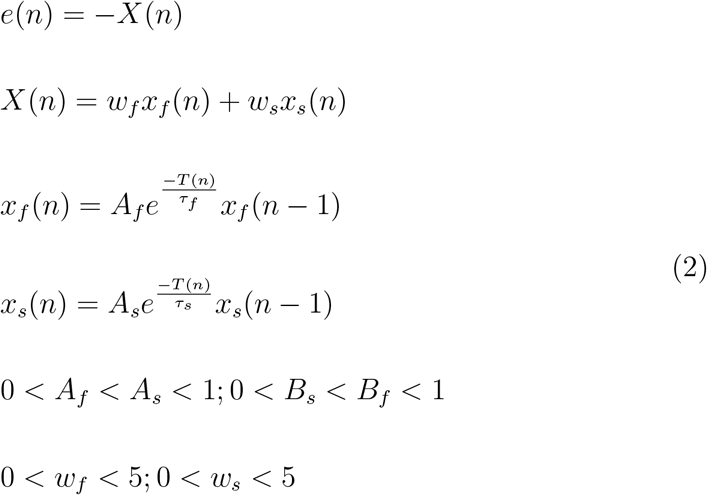

where *w*_*f*_ and *w*_*s*_ are weighting factors that introduce a dissociation between the state of a given system on prism exposure blocks and the total amount of learning by each system that is expressed in after-effect blocks. With these free parameters the after-effect is now modelled as a weighted sum of the states of the two systems. For each system, having a weight lower than 1 means a downscaling of the learning by that system that is expressed in the after-effect, whereas a weight greater than 1 means an over-expression.

The model was fitted using a least squares procedure with the *fmincon* function in MATLAB (The MathWorks inc., version 2014b). Because of the MRI scanning environment, obtaining a reliable measure of endpoint errors posed a challenge, particularly for stroke patients (Bultitude et al., 2016). As a result, individual behavioural datasets were noisier than those typically collected outside the scanner. This prevented us from obtaining separate model parameter estimates for each participant. To overcome this issue, the weighted two-state model was fitted to the averaged pointing data of the healthy control group. Individual time courses of the fast and slow system dynamics were subsequently generated for each participant using participant-specific event onset information. The use of control participant dynamics to model learning and forgetting in stroke patients was validated using behavioural data collected outside the scanner. That is, we compared model computational parameters between healthy adults and neglect patients, showing similar estimates for both groups (see Appendix .2).

#### 2.4.3. Modelling fast and slow systems using computational fMRI

Two computational time-course regressors, representing the states of the fast and slow system, were extracted from the group averaged behavioural data and yoked to each participant’s (healthy control and stroke patient) fMRI event timings for the real prism adaptation run. The values for the fast and slow system states were yoked to two time-points for each trial: a)the presentation of the pointing target, and b) when participants touched the presentation screen. That is, the state of the two systems was sampled twice per trial to capture two key time points within each trial: a) movement preparation, and b) error feedback. The FMRIB Software Library v6.0 was used for processing and analysis of fMRI data (Jenkinson et al., 2012).

A lesion mask was manually created for each stroke patient from the structural scan using FSLview software under the direction of a neurologist. Lesion masks were used to optimise image registration for stroke patients. The fMRI pre-processing steps comprised: a) brain extraction, b) geometrical distortion correction using participant-specific field maps, and c) denoising using Independent Components Analysis (ICA). Seven explanatory variables were used to model blood-oxygen-level dependent (BOLD) activity during sham and real prism runs using FSL’s FEAT toolbox (Woolrich et al., 2001, 2004): 1) the timing of each target presentation and screen touch during error correction trials, modelled as a constant ‘stick’ function, 2) the timing of each target presentation and screen touch during after effect trials, modelled as a constant ‘stick’ function, 3) a linear trend spanning the length of the fMRI run, ranging from 1 to −1 to account for linear habituation effect in BOLD signal owing to repeated pointing over time, 4) the value of the fast system during error correction trials, 5) the value of the fast system during after-effect trials, 6) the value of the slow system during error correction trials, and 7) the value of the slow system during fter-effect trials. A task-average prethreshold mask was applied to increase analysis sensitivity, defined as regions activated by the group average activation map across sham and real prism adaptation runs, constrained to gray matter regions (refer to Appendix .3 for images of the fMRI EVs and pre-threshold masks). Group effects were estimated using FSL’s Flame 1 + 2 with outlier detection. The main contrasts of interest were the average activation maps for the fast and slow systems during the real prism adaptation runs. Significant activation clusters during the real prism run for the fast and slow computational regressors were interrogated by comparing percent signal change between sham and prism runs using binarised cluster masks. These contrasts were conducted using one-tailed paired-samples t-tests [prism>sham].

## 3. Results

The primary goal of the study was to identify brain regions where the time-course of functional activation was correlated with the gradual stabilization of the prism after-effect, represented in the computational modeling as the timecourse of the slow system. The fast and slow time-courses were modelled based on the age-matched healthy control behavioural data and applied to both healthy and stroke participants. Brain regions displaying significant activation to fast or slow system regressors were further interrogated by comparing percent signal change between sham and real prism adaptation runs.

### 3.1. Behaviour: real, compared to sham, prism adaptation results in a left-ward pointing bias in after effect trials

Between-group (control vs stroke) and within session (sham vs real) differences in leftward pointing errors were evaluated using a linear mixed effect model (Figure 1). The real prism adaptation run resulted in significantly greater leftward error deviation (*M* = − 43.6 pixels, *SD* = 45.58) compared to the sham run (*M* = .7 pixels, *SD* = 63.32) (Run parameter estimate = − 40.4, 95% CI [− 45.24, − 35.64], *p <* .001). There was no difference in leftward after effect magnitude between healthy controls (*M* = − 26.0 pixels, *SD* = 57.1) and stroke participants (*M* = −13.1 pixels, *SD* = 62.7), indicating similar behavioural adaptation across the two groups (Group parameter estimate = −1.1, 95% CI [− 24.86, 22.42], *p* = .92). There was also no interaction between group and run (Group x Run parameter estimate = − 3.3, 95% CI [− 9.36, − 2.68], *p* = .276). That is, the magnitude of left-ward after effects did not differ between groups for the sham (Control: *M* = − 4.4 pixels, *SD* = 61.61, Stroke: *M* = 8.9 pixels, *SD* = 65.20) or real (Control: *M* = − 48.0 pixels, *SD* = 41.81, Stroke: *M* = − 36.29 pixels, *SD* = 50.42) fMRI runs.

**Figure 1:**
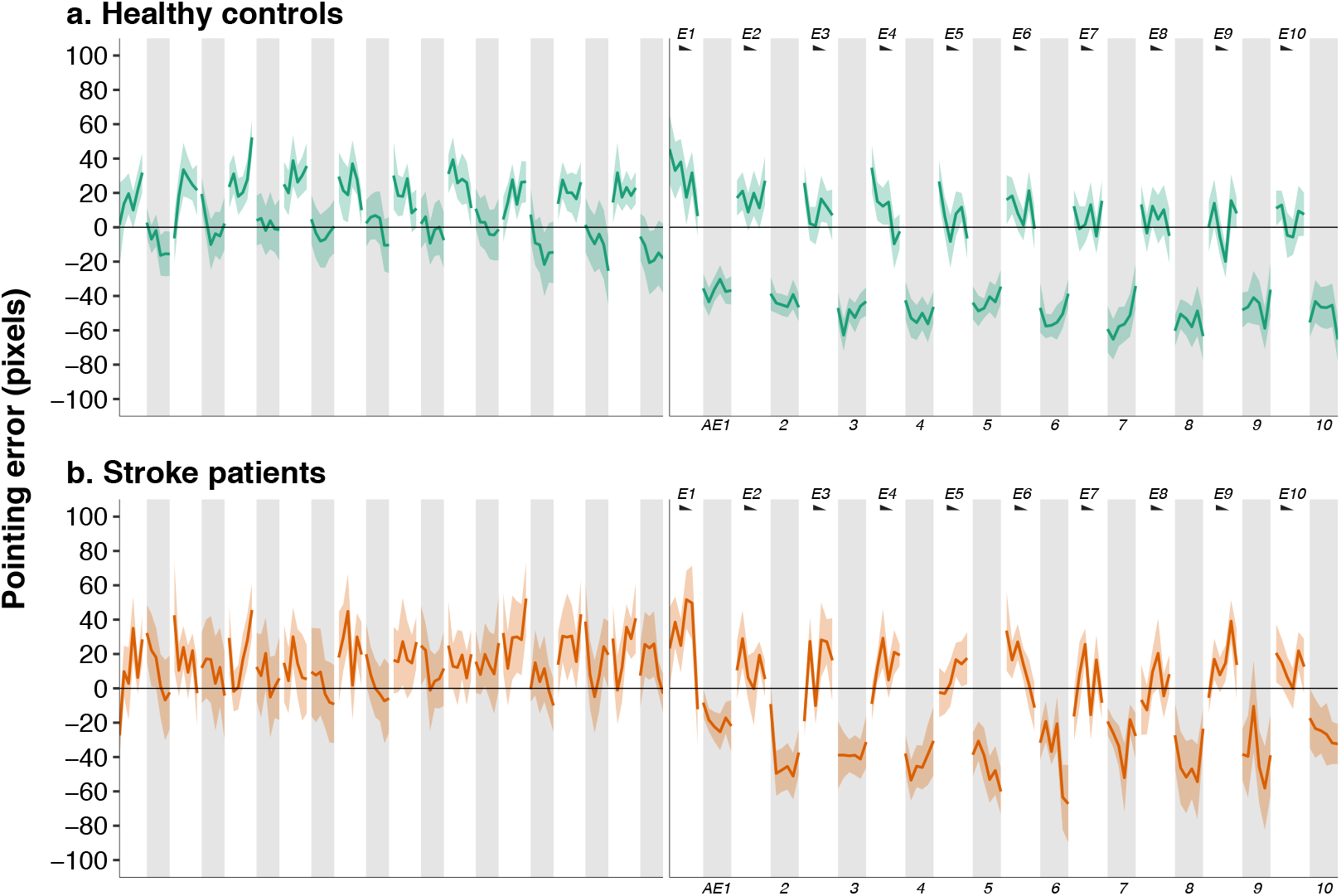
Prism adaptation results in an after-effect in both healthy controls and stroke patients. The figure shows trial-by-trial pointing errors in the sham run (left panel) followed by the prism run (right panel). Blocks E1-16 show error correction trials during prism exposure. Interleaved blocks (AE1-10) show blocks of after-effect measurement. During the AE blocks there was significantly greater leftward pointing errors (after-effects) during real, compared to sham, prism adaptation. **a**. Trial-by-trial pointing errors by healthy controls (n = 17). **b**. Trial-by-trial pointing errors by stroke patients (n = 11). Prism exposure blocks (white columns) and after effect blocks (grey shaded columns) show trials averaged across participants together with +/-1 standard error of the mean error bras (ribbon shading). A linear mixed-effect regression indicated a significant main effect of Run [prism vs sham] (Run parmeter estimate = − 40.4, 95% CI [− 45.24, −35.64], *p <* .001), but no effect of Group [patients vs controls] (Group parameter estimate = − 1.1, 95% CI [−24.86, 22.42], *p* = .92) and no Run x Group interaction effect (Group x Run parameter estimate = −3.3, 95% CI [− 9.36, − 2.68], *p* = .276). Thus patients and healthy controls showed behavioural adaptation to prisms that differed from behaviour during the sham run but that did not differ between the groups **E** = Prism exposure block, **AE** = After-effect block

### 3.2. A weighted two-state computational model accounts for prism adaptation behaviour

A weighted two-state computational model was fitted to the averaged real prism adaptation behavioural data of healthy control participants (Figure 2, panel A). The model showed a good fit to the data (*R*^2^ = .93, *RMSE* = 8.279). This model displays predicted behaviour: a) fast system dynamics (red) display rightward errors at prism onset that are corrected quickly across subsequent trials; b) slow system dynamics (blue) increasingly dominate as prism after-effects stabilize across trials. Owing to a high amount of noise in stroke patients’ in-scanner pointing, fitting the model using the stroke patient pointing data resulted in a poor model fit *R*^2^ = .74, *RMSE* = 14.328. This poor fit is indicated by: a) the fast system (red) dominating at all stages during the prism adaptation procedure; b) the weight of the slow system estimated to be 0 during after-effect trials. Because of this the model fit (purple) during after-effect trials is equivalent to the value of the fast system, rather than being a weighted averaged of both systems. Given the poor fit of the model using stroke participant data, instead we inferred the internal state dynamics of stroke patients using the computational model developed on control data. To validate this approach, we compared pointing dynamics outside the scanner, which confirmed behavioural similarity between healthy and stroke cohorts (see Appendix .2),

**Figure 2:**
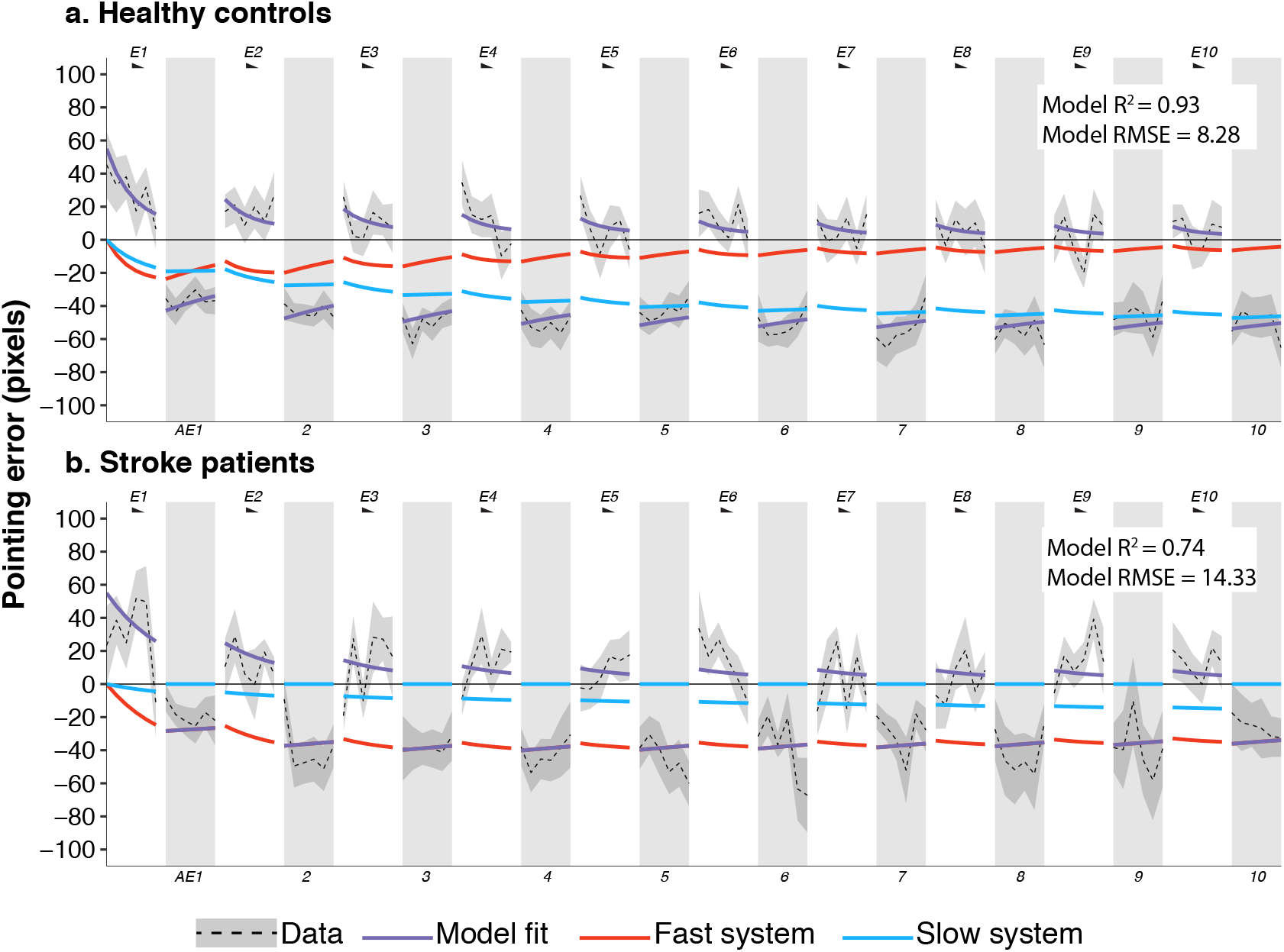
Fitting the weighted two-state computational model to prism adaptation behaviour. This figure shows a good fit of the weighted two-state model to healthy control pointing dynamics over time (panel a), but poor fit to stroke patients (panel b). **a**. The overall *R*^2^ = .93 (higher is better) and *RMSE* = 8.28 (lower is better), measures of the overall model fit, show a good fit to the healthy control data. The overall model fit (purple) is the sum of two underlying computational systems: a fast system (red) that is expressed maximally early in the adaptation process and gradually dissipates, and a slow system (blue) that increases in magnitude throughout adaptation as the after effect stabilizes over time. Parameters from this model were used for subsequent control and stroke participant fMRI analyses. **b**. The overall *R*^2^ = .74 and *RMSE* = 14.33 indicate a poorer fit to stroke patient pointing dynamics. Poor model fit is also illustrated in the following ways: a) the fast system (red) is dominant at all stages during the prism adaptation procedure; b) the weight of the slow system is estimated to be 0 during after effect trials.

### 3.3. Prism adaptation after-effect stabilization in sensorimotor cortex

The fast and slow computational time-courses were fitted to prism adaptation behaviour and regressed against BOLD fMRI activity. The primary contrasts of interest were the average activation maps for the fast and slow system timecourses during the error correction and after effect blocks of the real prism adaptation run. During the real prism adaptation run there were no significant activations for the fast system time-course, neither for control nor for stroke patients. However, for healthy controls the slow system time-course correlated with BOLD activity in a single left sensorimotor cortex cluster (Figure 3, panel A (red); cluster information: 136 voxels, p = .004, peak Z = 4.26, peak MNI coordinates = [x: −34, y: −38, z: 70]). This region showed greater percent signal change during the real (*M* = .55%, *SD* = .59, 95% CI [.27, .83]) compared to sham (*M* = .13%, *SD* = .24, 95% CI [.02, .25]) prism adaptation run (Figure 3, panel A right; *t*(16) = 2.59, *p* = .01, *one* − *tailed*). For participants with stroke, the slow system time-course also correlated with BOLD activity in left sensorimotor cortex (Figure 3, panel B (red); cluster information: 98 voxels, p = .002, peak Z = 4.05, peak MNI coordinates = [x: −24, y: −28, z: 60]) as well as in the right cerebellum (Figure 3, panel C (red); cluster information: 111 voxels, p = .001, peak Z = 4.94, peak MNI coordinates = [x: 8, y: −52, z: −12]). The sensorimotor cluster showed greater percent BOLD signal change during the real (*M* = .82%, *SD* = .95, 95% CI [.26, 1.38]) compared to sham (*M* = .24%, *SD* = .21, 95% CI [.12, .36]) prism run (Figure 3, panel B; *t*(10) = 1.83, *p* = .048, *one* − *tailed*). There was no difference in percent BOLD signal change between the real (*M* = .69, *SD* = .39, 95% CI [.46, .92]) and sham(*M* = .52, *SD* = .96, 95% CI [− .05, 1.08]) runs for the cerebellar cluster (Figure 3, panel C right; *t*(10) = .52, *p* = .31, *one* − *tailed*).

**Figure 3:**
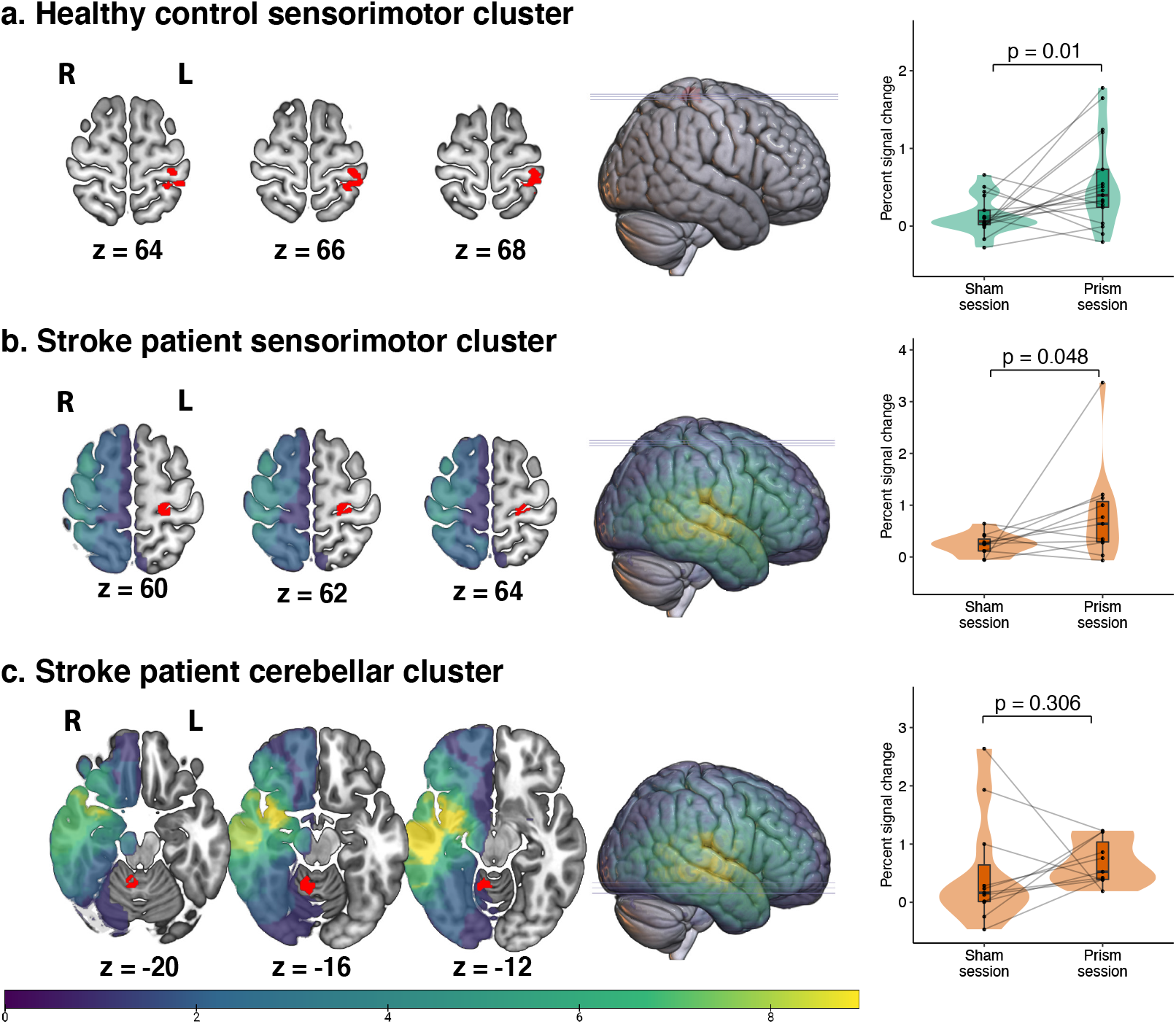
Sensorimotor cortex correlates with stabilization of the prism adaptation after effect in healthy controls and stroke patients. The figure shows BOLD activity clusters that correlate significantly with the slow system computational timecourse for healthy controls and stroke patients. **a**. Significant activation cluster (red) for the slow system time-course for healthy controls. A single activation cluster was found in left sensorimotor cortex. Panel a, right compared percent signal change between the sham and real prism adaption runs. Violin plots illustrate the distribution of signal within each run, and are overlaid by boxplots and individual participant datapoints. The sensorimotor cluster demonstrated significantly greater percent signal change in the real prism run compared to the sham run (*t*(16) = 2.59, *p* = .01, *one* − *tailed*) **b**. Significant sensorimotor activation cluster (red) for the slow system time-course for stroke patients (Frequency lesion mask is overlaid; brighter colours = greater lesion overlap across patients). Panel b, right compared percent signal change between the sham and real prism adaption runs. The sensorimotor cluster demonstrated significantly greater percent signal change in the real prism run compared to the sham session (*t*(10) = 1.83, *p* = .048, *one* − *tailed*). **c**. Significant cerebellar activation cluster (red) for the slow system time-course for stroke patients. Panel c, right compared percent signal change between the sham and real prism adaption runs. There was no difference in percent signal change between real and sham runs *t*(10) = .52, *p* = .31, *one* − *tailed*).

**Figure 4:**
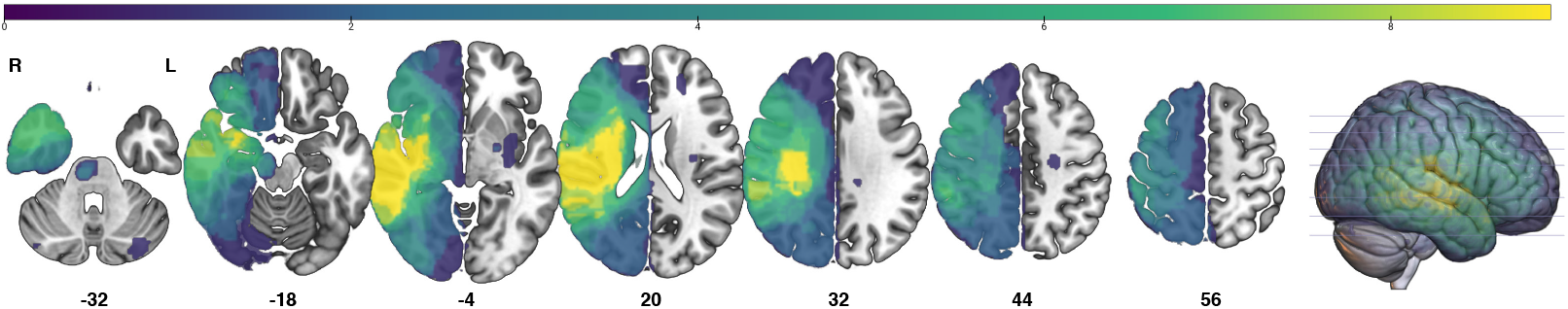
Stroke patient lesion overlay image. This figure shows lesion overlay maps for all stroke patients. Bright colour indicates greater overlap.

## 4. Discussion

In this study, we aimed to identify the brain circuits in which functional activity tracks the development and stabilization of the prism adaptation after effect. This is a critical gap in the field, given that the consolidation, and persistence, of the after-effect is arguably the key behaviour that drives the therapeutic response to prism adaptation in neglect rehabilitation (Jacquin-Courtois et al., 2013, O’Shea et al., 2017). To do so, we leveraged computational modelling approaches to prism adaptation (Körding, 2007, Joiner and Smith, 2008, Petitet et al., 2018), in conjunction with fMRI. This approach enabled us to identify a slowly evolving computational timecourse during prism adaptation that correlates with activity in sensorimotor cortex and gradual stabilization of the prism after effect. The same region was activated both in healthy controls and stroke patients.

As summarised by Panico et al. (2019), only a small number of fMRI studies of prism adaptation have been performed (Clower et al., 1996, Danckert et al., 2008, Luauté et al., 2009, Chapman et al., 2010, Küper et al., 2014, Saj et al., 2013, 2019). In part, this is due to technical and design challenges, such as arm movement related artifacts (Bultitude et al., 2016). Studies to date have primarily focused on the initial phase of prism adaptation, which is dominated by strategic error correction (Clower et al., 1996, Danckert et al., 2008, Luauté et al., 2009), for example, by contrasting brain activity during the first few pointing trials with the last few trials. Such studies have identified cortical regions crucial for error detection and correction, including the posterior parietal cortex, anterior cingulate, intraparietal, and medial cerebellar regions. In the current study, we found no correlation between the fast computational timecourse–a putative measure of strategic error correction–and brain activity. It might be that the temporal resolution of fMRI hemodynamic responses are simply too slow to detect rapid changes associated with strategic error correction. Indeed, prior studies may have been able to detect brain activation related to strategic error correction by examining cumulative brain activity averaged across trials.

From a neuro-rehabilitation perspective, identifying the neural correlates of prism after effect formation and stabilization is arguably of greater relevance, due to the perceived importance of pointing after effects for knock-on therapeutic effects on cognition. Whilst inter-related, error correction and after effect consolidation are independent processes–the time-course of the latter cannot readily be inferred from the former (Michel et al., 2007, Newport and Jackson, 2006). Luauté et al. (2009) were the first to use a protocol that enabled the examination of brain activity beyond the initial error correction phase. Whereas parietal and occipital regions were active early in adaptation, the cerebellum and superior temporal regions were active during late stages when errors had been corrected back to baseline levels of accuracy. Subsequent studies have implicated similar, but also divergent, brain regions (Chapman et al., 2010, Küper et al., 2014, Saj et al., 2019). In a study with neglect patients, for example, the therapeutic effects were associated with activity of posterior parietal and mid-frontal cortex (Saj et al., 2013).

In contrast to these prior studies, the present work aimed to characterize the neural correlates of prism after effect formation and consolidation (Bultitude et al., 2016). Instead of the traditional three-stage prism adaptation paradigm comprising baseline, exposure, and post-exposure, we interleaved blocks throughout the prism adaptation procedure that captured the development of an after-effect by removing visual feedback of pointing movements. Using a computational neuroimaging framework, we were able to identify for the first time that trial-by-trial activity in sensorimotor cortex tracks the slow system process of after effect consolidation over time. One of the limitations of contrasting early vs late pointing trials is the inability to disentangle the contribution of multiple cognitive processes that coexist in time. That is, signal in the early and late stages are a combination of multiple processes. Because the time-course of different processes are not explicitly modelled in such approaches, it is similarly difficult to establish the temporal course of brain activation at different stages. For example, a region may be deactivated early, returning to its normal level of activation in late stages, whilst another region may become activated only in a later stage.

Only one other recent fMRI study has used a similar analytic approach– extracting internal system states using computational modelling and regressing those behavioral signatures against fMRI brain activity (Kim et al., 2015). That study found that fast internal states correlated with activity in frontal and parietal cortices as well as posterior-lateral cerebellum, intermediate internal states with the inferior parietal lobe, and slow states with the anterior-medial cerebellum. The findings of the current study, implicating sensorimotor cortex, differ from those reported by Kim et al. (2015), likely due to key differences in study methodology. Kim et al. (2015) employed a visuomotor rotation task requiring participants to adapt to two opposing visual perturbations using small joystick movements. Visual feedback of performance was provided by the position of a dot on a screen. This is in contrast to our prism adaptation paradigm that requires participants to adapt to, and consolidate, a single visual perturbation using hand movements. Visual feedback was of the participant’s own hand position at the end of the pointing movement. That is, these two studies differed both in terms of *type of movement* and *visual feedback*. Whilst prism adaptation has consistently been shown to induce a proprioceptive after-effect–a change in the experience of the limb in space–it is unclear whether a similar after-effect is induced by the visual rotation paradigm employed by Kim et al. (2015), Facchin et al. (2019), using a joystick rather than whole-arm movements. Differences between the studies might also relate to the way in which individuals assign the ‘credit’, or reason, for their performance errors (Kording et al., 2007). That is, in the joystick-task individual’s may have learned more about how to manipulate the tool, rather than their own body, to solve the problem (Kluzik et al., 2008, Yamamoto et al., 2006), consequently forming an internal model of the tool (joystick) rather than of their own arm movements (Fleury et al., 2019).

Involvement of sensorimotor cortex in adaptation, learning, and skill acquisition is supported in a number of studies (Muellbacher et al., 2002, Landi et al., 2011, Hadipour-Niktarash et al., 2007, Ogawa and Imamizu, 2013, Paz et al., 2005). For example, by employing a visuomotor rotation task in monkeys, Paz et al. (2005) found that dynamic reorganisation of distinct cortical areas took place at different time-points during adaptation. Whereas supplementary motor areas mediated the early stage of adaptation, changes in the primary motor cortex were observed at only the late stage of adaptation. As interpreted by the authors, the late activation in the primary motor cortex could reflect the shift of the new acquired skill from working- to long-term memory. Thus, it is possible that the critical role of the sensorimotor cortex may have gone unnoticed in previous prism adaptation studies due to lack of a clear timecourse representing motor memory consolidation, and the paradigms themselves did not sufficiently elicit the need for brain circuitry crucial for longer-term consolidation, instead stressing strategic error detection and correction mediated by cerebellar, parietal, and frontal cortical regions.

One major limitation of this study was the reliance on a computational model that was developed solely on healthy control pointing movements. That is, due to unacceptable levels of noise in pointing dynamics for stroke patients, the model parameters that best fit healthy controls were used to fit the fMRI data both healthy and stroke patients. Although we confirmed similar prism adaptation pointing behaviour and comparable model parameters out-of-scanner for healthy and stroke patients, the results of the current study need to be interpreted within this constraint. That is, our findings should be interpreted with regards to similarities, rather than differences, between healthy controls and stroke patients. Specifically, in this study we were only able to ascertain whether similar brain circuitry encoded a latent memory trace that represents consolidation. Encouragingly, we show that the same behavioural computation is mediated by similar brain circuitry. This is especially relevant to experimental studies in healthy individuals that may more readily translate or generalise their findings to stroke populations.

Despite the translational relevance of the current findings, another important study limitation was that the majority of stroke patients were not presenting with symptoms of visuospatial neglect at the time of study participation. Therefore, we do not know to what extent the sensorimotor regions identified in the study relate to presence or severity of attentional neglect, or whether activity in the sensorimotor region identified in the current study is causally implicated in the therapeutic effect of prism adaptation. Previous work has shown that noninvasive brain stimulation of sensorimotor cortex can prolong retention of the prism after-effect (O’Shea et al., 2017). Future studies can strengthen current findings by having clinical pre-post measures following prism adaptation and examining how neurostimulation of sensorimotor cortex alters slow system dynamics (and consequently the therapeutic effect).

This paper moves the field forward by identifying sensorimotor cortex as a key brain region mediating the consolidation of prism after effects. This is useful for future studies attempting to increase the longevity of the therapeutic effects of prism adaptation. Enhancing/optimising sensorimotor cortex functional activation in this may lead to greater reductions in spatial neglect following stroke.

## Data Availability

Data will be made available upon reasonable request.

## Appendix .1. Stroke patient lesion overlay image

## Appendix .2. Comparing out-of-scanner prism adaptation dynamics between healthy controls and neglect patients

We employed two approaches to ensure the validity of applying the weighted two-state model parameters based on healthy control dynamics to stroke patients: a) we compared the dynamics of prism adaptation both visually (observed pointing dynamics) and, b) quantitatively (i.e. statistically comparing individual computational model parameter estimates) of historical behavioural data collected outside the scanner. We used data from 34 healthy controls (mean age = 66.68, SD = 8.44) and 15 neglect patients (mean age = 59.24, SD = 7.78). From a qualitative perspective, Figure .5 indicates similar pointing dynamics between healthy older adults and neglect patients.

**Figure 5:**
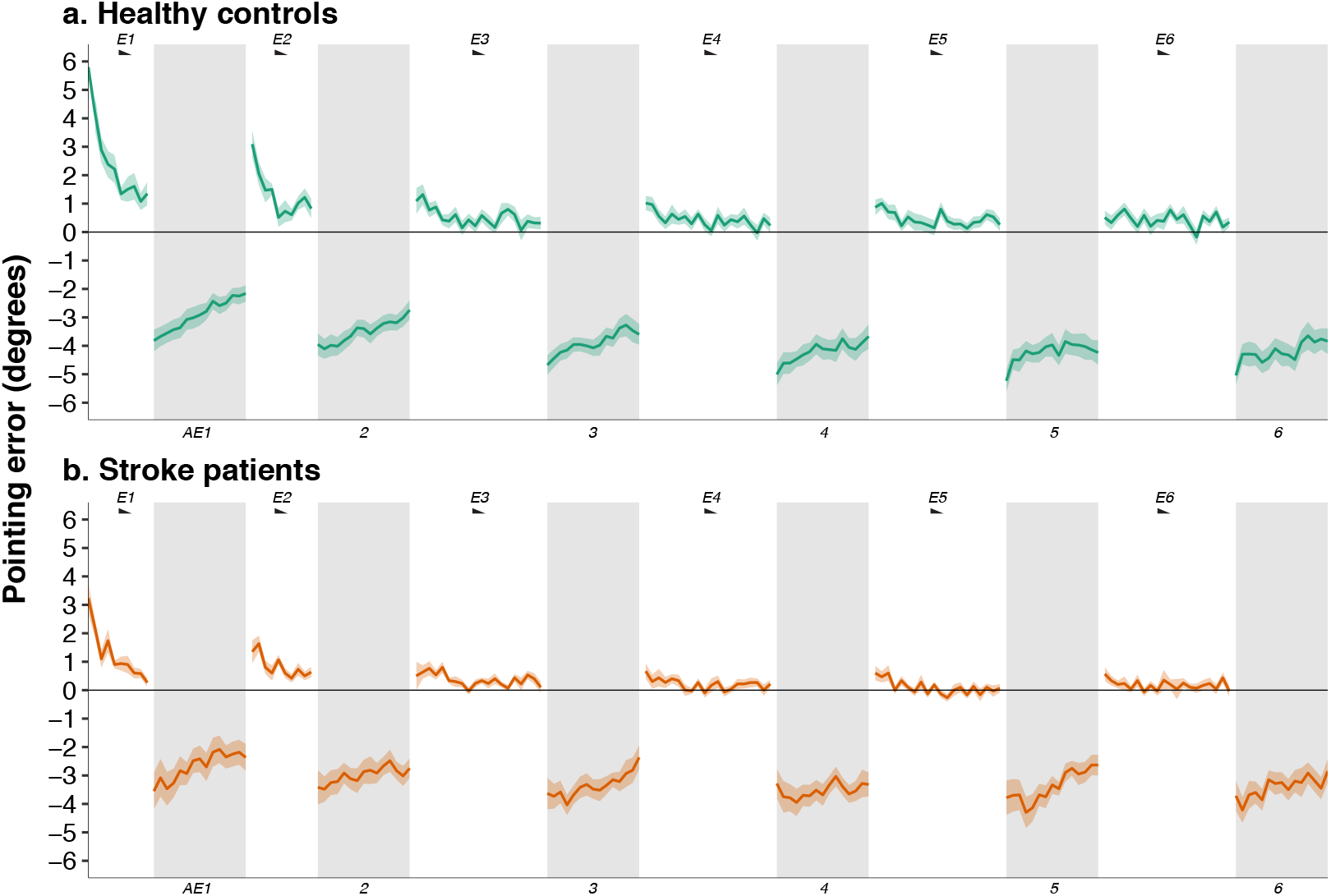
Pointing dynamics for healthy older adults and stroke patients collected out-of-scanner. This figure shows the pointing dynamics from a prism adaptation protocol collected in a non-MRI environment, where measurement noise is significantly reduced. It can be seen that healthy older adults (green) and neglect patients (orange-brown), in general, display similar pointing dynamics for AE (white columns) and error-correction (grey shaded columns) trials.

**Figure 6:**
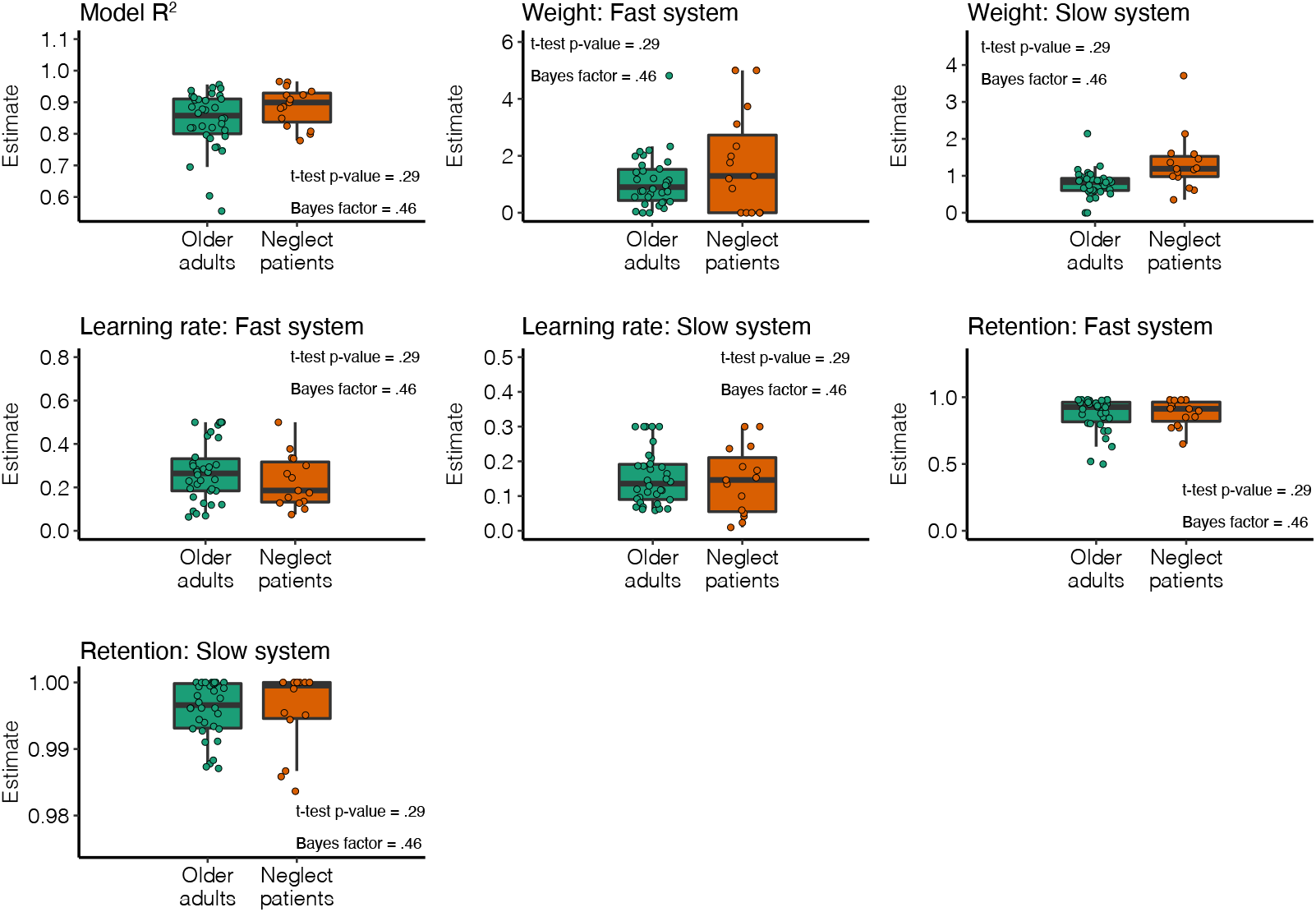
Model parameter comparison. This figure shows comparisons of model computational parameters between healthy older adults and neglect patients. p-values following two-tailed independent samples t-test are presented as well as Bayes factor values. Bayes factor presents the strength of evidence for a particular hypothesis. A Bayes factor between1/3.2 and 3.2 indicate no evidence for the hypothesis.

In contrast to inside the scanner, the precise timing of each trial was un-known. Thus, similar to previous studies (Inoue et al., 2015, Joiner and Smith, 2008, Smith et al., 2006), retention factors were defined as a function of trial rather than time. The model was therefore defined as follows on error-correction trials:

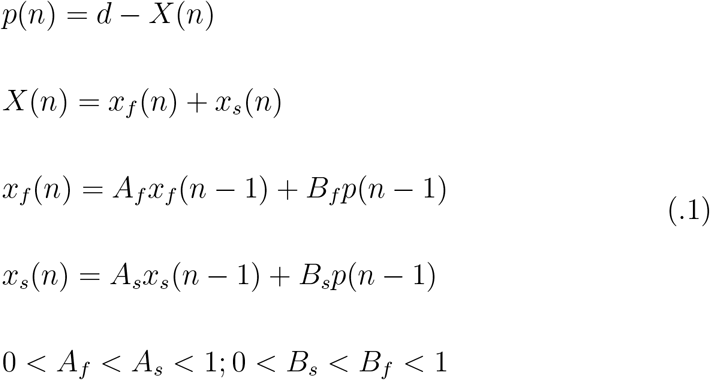

And as follows on AE trials:

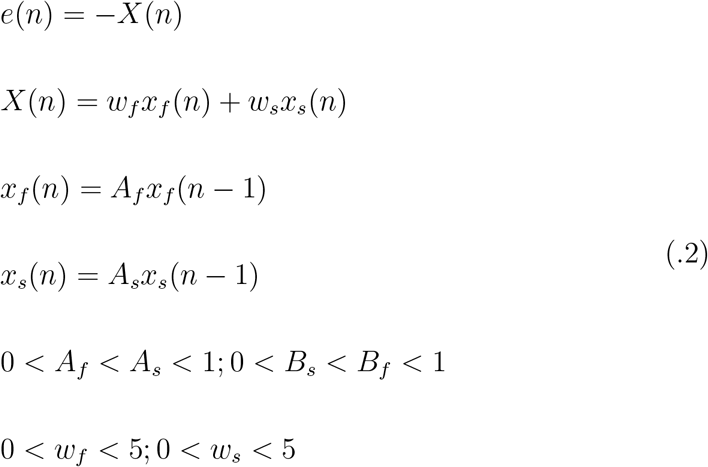

The model differs in four ways between the two pointing types. First, a different bias term is used, reflecting the fact that, at baseline, individuals show different biases depending on visual feedback availability. This bias was computed as the average endpoint error for each type of trial during the pre-exposure phase. Second, the prism effect *d* is set to zero on AE trials (no prism deviation). On error-correction trials, *d* is set to the group-averaged normalised pointing error obtained on the first exposure trial (i.e. when no adaptation has occurred yet). This corresponded to *d* = 3.33*cm* for the neglect group, and *d* = 6.07*cm* for the control group. Third, the learning rates of the fast and slow systems are set to zero on OLP trials in order to reflect the absence of visual feedback. Fourth, weighting factors are introduced on OLP trials to allow for a dissociation between the state of a system on CLP and its contribution to the after-effect on OLP.

After optimising the model to individual datasets (6 free parameters), the quality of the fit was comparable across the two groups (r-square; (*t*(39.6) = 2.01, *p* = .051, *two* − *tailed*)). We found no evidence of a difference in learning (fast system: *t*(30.3) = − 1.08, *p* = .290, *two* − *tailed*; slow system: *t*(22.8) = − .35, *p* = .727, *two* − *tailed*) or forgetting (fast system: *t*(34.8) = .24, *p* = .814, *two* − *tailed*; slow system: *t*(17.4) = − .76, *p* = .458, *two* − *tailed*) between patients and controls. This suggested that similar learning and forgetting dynamics were driving overt behaviour during prism adaptation in patients and controls.

However, the two groups differed in terms of the weight attributed to the slow system during OLP trials (*t*(16.8) = 2.56, *p* = .020, *two* − *tailed*), with patients showing greater transfer. The contribution of the fast system to the after-effect on the other hand was not significantly different between the two groups (*t*(17.60) = 1.34, *p* = .20, *two* − *tailed*). The apparent over-weighting of the slow system reflected the fact that patients showed after-effects that were overall comparable to controls, despite making smaller pointing errors during exposure trials. In our experience, smaller endpoint errors during prism exposure result from the fact that stroke patients tend to move slower, which allows them to correct their movement in-flight to a greater extent than controls. Thus, it might be the case that overt pointing behaviour better reflects the error signal driving adaptation in controls, whereas it is contaminated by online corrections in stroke patients.

In order to test this idea and investigate whether it accounted for the apparent over-weighting of the slow system in stroke patients, we introduced an *overt expression* parameter on CLP trials as follows:

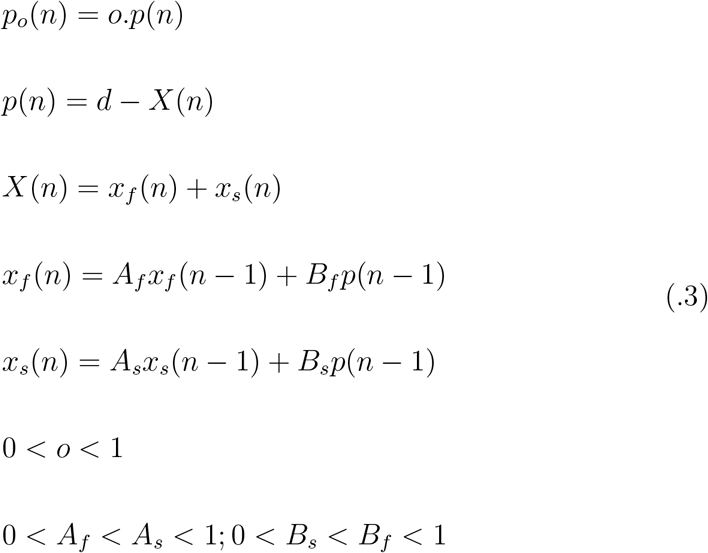

where *o* expresses the fraction of error signal that is expressed in overt behaviour after online correction. A value of zero means perfect online control (none of the prism-induced error is expressed in the endpoint error), while a value of one means no online control (all of the prism-induced error is expressed in the endpoint error). Crucially, the systems learn based on the covert error, i.e. the error before online correction. This modification also allowed us to use the same prism effect *d* for the two groups, which guaranties a better comparison of the dynamics obtained.

As expected, this parameterisation of the model captured a difference of online control between the two groups. However, none of the other free parameters were significantly different. This reinforced the idea that the fast and slow adaptation dynamics were comparable across the two group, despite patients displaying smaller CLP errors overall, potentially due to larger online control.

## Appendix .3. fMRI modelling materials

**Figure 7:**
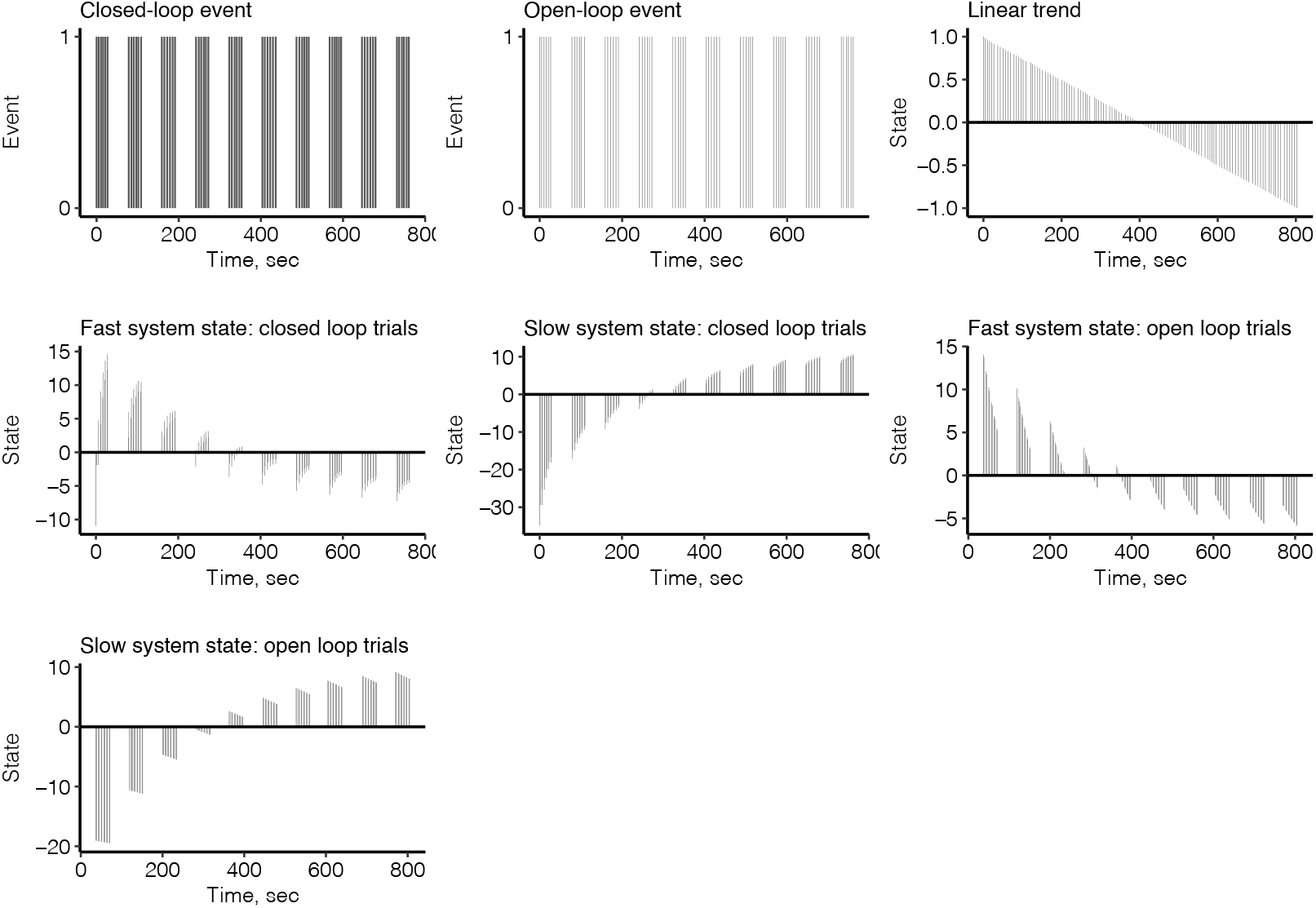
FMRI explanatory variables. The seven explanatory variables entered into the fmri general linear model.

**Figure 8:**
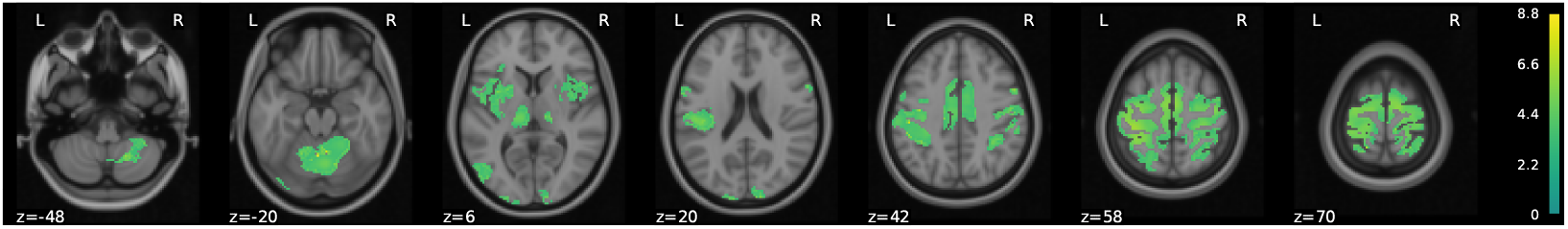
Task relevant mask for healthy controls. The ‘task-relevant’ mask used for control participants prior to statistical thresholding.

**Figure 9:**
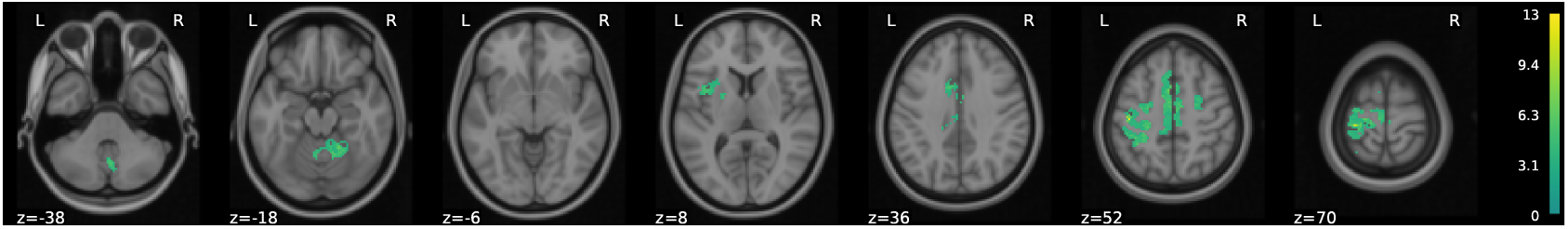
Task relevant mask for stroke patients. The ‘task-relevant’ mask used for control participants prior to statistical thresholding.

